# Prodromal manifestations of Parkinson’s disease in adults with 22q11.2 microdeletion syndrome

**DOI:** 10.1101/2022.05.18.22275282

**Authors:** Carlos Juri, Pedro Chaná-Cuevas, Vasko Kramer, Rosemarie Fritsch, Claudia Ornstein, Analía Cuiza, Carlos Hernández, Katiuska Villanueva, Teresa Cordova, Jorge Mauro, Adrian Ocampo, Boris Rebolledo-Jaramillo, Gonzalo Encina, Andrea Calleja, Javiera Dinator, Juan Pablo Alcayaga, Nicolas A. Crossley, Gabriela M. Repetto

## Abstract

22q11.2 microdeletion syndrome (22qDS) was recently identified as a risk factor for development of early-onset Parkinson’s disease (PD). The classical motor manifestations of this disease are preceded by early signs and symptoms of neurodegeneration. The progression of 22qDS-associated PD is unknown. We aimed to evaluate the presence of prodromal PD in a group of adults with 22qDS using the Movement Disorders Society (MDS) Criteria for Prodromal PD. Thirty-eight persons with 22qDS and 13 age-matched controls participated in the study, and their results were compared using the Mann-Whitney U test. Persons with 22qDS had lower scores on olfaction testing (p=7.42E×10^−5^), higher scores on the COMPASS 31 scale for dysautonomia (p=2.28×10^−3^) and on the motor evaluation using Movement Disorder Society (MDS)-sponsored revision of Unified Parkinson’s Disease Rating Scale motor subscore (UPDRS-III) (p=1.84×10^−4^), compared with healthy controls. Home polysomnogram did not find participants with REM-sleep behavior disorder. Integrity of nigrostriatal dopaminergic system was evaluated by PET-CT imaging of presynaptic dopamine with ^18^F-PR04.MZ. Patients showed significantly higher specific binding ratios in the striatum, compared to controls (p=9.57×10^−3^ at the caudate nuclei). Two patients with 22qDS (5.2%) had decreased uptake in the posterior putamen (less than 60% of controls) and one fulfilled MDS criteria for prodromal PD. These results show that patients with 22qDS manifest some signs and symptoms of prodromal PD such as hyposmia, dysautonomia and mild movement alterations. In the majority, this was associated with elevated dopaminergic signaling, suggesting that loss of dopaminergic neurons may not be the cause. A smaller subgroup did show evidence of a decrease in nigrostriatal dopaminergic signaling, as seen in classical prodromal PD. Longitudinal studies are necessary to understand the progression to and risk of PD in persons with 22qDS.

## Introduction

22q11.2 microdeletion syndrome (22qDS) is the most common microdeletion syndrome in humans, with an estimated incidence of 1/2,200 to 1/10,000 live births (Blagojevic et al., 2021; Oskarsdóttir et al., 2004; Panamonta et al., 2016; Rehman et al., 2015). It causes major congenital anomalies, neurodevelopmental disabilities, and increased risk of schizophrenia (reviewed in (McDonald-McGinn et al., 2015)). Longitudinal follow up of cohorts into adulthood has led to the recognition that 22qDS is also a risk factor for early-onset Parkinson’s disease (EOPD). Persons with 22qDS-associated PD have clinical characteristics, therapeutic response, neuro-imaging and neuropathological findings of multifactorial/idiopathic Parkinson’s disease (PD), with an average age of onset of approximately 40 years, and an increase in risk estimated close to 70-fold compared to population risk at that age (Booij et al., 2010; Butcher et al., 2013, 2017b; Pollard et al., 2016; Zaleski et al., 2009). In addition, genome wide association studies identified 8 individuals with 22qDS in a large, unselected population of 9387 persons with presumptive “idiopathic” PD, with a median age of onset of 37 years (Mok et al., 2016).

The classic motor manifestations of PD, namely, bradykinesia, rigidity and tremor are preceded by about a decade by prodromal manifestations (Postuma et al., 2015; Postuma, Lang, et al., 2012). This early stage of PD heralds the onset of the classic motor features and is the result of gradual neurodegeneration (Braak et al., 2003, 2006). Prodromal PD is characterized by olfactory loss, dysautonomia, rapid eye movement (REM) behavior disorder (RBD) and subtle motor findings as well as gradual dopaminergic loss as evidenced by neuroimaging using different dopamine (DA) transporter (DAT) tracers (Berg et al., 2015). There is marked interest in studying prodromal PD and identifying subjects at this stage, since it may provide a window of opportunity for neuroprotection.

Recent studies show that some adults with 22qDS manifest some features suggestive of prodromal PD, such as hyposmia and motor abnormalities, but these have been assessed in non-PD related contexts or using selected prodromal markers. For example, Sobin et al (Sobin et al., 2006), and Romanos et al (Romanos et al., 2011) described increase frequency of olfactory dysfunction in children, adolescents and young adults with 22qDS, in the setting of evaluation of dopaminergic dysfunction as marker for psychosis risk. With the aim of assessing prodromal PD features in adults with 22qDS, Buckley et al(Buckley et al., 2017) found increased rates of hyposmia, and symptoms suggestive of dysautonomia and RBD, using questionnaires in a cross-sectional study of 50 adults, and also found subtle motor alterations, suggestive of parkinsonism, in 4 of them. Furthermore, Butcher et al (Butcher et al., 2017b) reported similar findings of increased rates of hyposmia in 13 adults without and one with PD, and included neuroimaging markers to assess the risk of PD such as transcranial sonography and evaluated dopaminergic loss by positron emission tomography (PET/CT) with ^11^C-DTBZ, a radioligand that binds to the presynaptic vesicular monoamine transporter type 2. These studies found the expected reduced nigrostriatal PET/CT signaling in the patient with 22qDS associated PD, but found significantly increased hyperechogenicity in the area of substantia nigra and elevated striatal ^11^C-DTBZ binding in persons with 22qDS without PD, when compared with age-matched healthy controls. These cross-sectional studies indicate the presence of manifestations similar to prodromal PD, and suggest that mechanisms underlying these findings in some patients may not be clearly related to a decrease in dopaminergic activity.

In this study, we aimed to systematically assess the presence of prodromal PD in a group of adults with 22q11DS at high risk for PD, using the Movement Disorders Society Research Criteria ^8^, that includes additional markers and an integrated risk estimation for progression to PD.

## Methods

This was a cross-sectional, case-control study in Chilean adults with 22q11DS. Molecular diagnosis was confirmed by FISH or MLPA. Control subjects were a group of healthy volunteers, of similar age range. The study was approved by the Ethics Committee at Facultad de Medicina, Clínica Alemana Universidad del Desarrollo, and participants and/or guardians gave written informed consent. The evaluations were carried out between June 2017 and October 2019.

For cases, clinical information regarding 22qDS manifestations were obtained from medical records and interviews of patients and/or guardians. All underwent psychiatric evaluation using the Spanish versions of the Mini-International Neuropsychiatric Interview (MINI) version 5.0 (Ferrando et al., 1998), and cognitive evaluation the Wechsler Adult Intelligence Scale (WAIS-IV) (Rosas et al., 2014). Whole-night home polysomnograms were performed by means of a portable Type-2 PSG device (Alice PDX Diagnostic Systems, Phillips Respironics, Murisvile, PA) during habitual bedtime, and results scored using the American Society of Sleep Medicine (AASM) (2007) (Frauscher & Högl, 2015; Iber et al., 2007, Mauro et al., 2022). Search for pathogenic variants in a panel of 24 known genes for monogenic PD genes was done in 33 participants via next generation sequencing. Pathogenicity of identified variants was estimated using Combined Annotation Dependent Deletion (CADD) score >15 (Kircher et al., 2014). Genome Aggregation Database (gnomAD) database was used as a reference for population allelic frequency (Genome Aggregation Database (gnomAD). The list of sequenced genes can be found in SupplementaryTable 1.

All participants, cases and controls, underwent a complete neurological examination, including the Movement Disorder Society-Sponsored revision of Unified Parkinson’s Disease Rating Scale motor subscore (UPDRS-III). (Goetz et al., 2008). Parkinsonism was diagnosed when subjects presented bradykinesia and rigidity, with or without tremor. Olfaction was evaluated with Sniffin’ Sticks (Burghart Instruments, Wedel, Germany). Dysautonomia was evaluated using the Composite Autonomic Symptom Score (COMPASS31) scale (Sletten et al., 2012)-The RBD-13 and the RBD-1Q screening questionnaires were answered by participants or caretakers living with them(Postuma, Arnulf, et al., 2012; Stiasny-Kolster et al., 2007). Integrity of dopaminergic neurons in nigrostriatal regions was evaluated by PET/CT imaging of DAT with ^18^F-PR04.MZ, as previously described (Juri et al., 2021; Kramer et al., 2020; Lehnert et al., 2022).

Results from these evaluations were stored in RedCap, (Harris et al., 2009). Descriptive analysis was performed using median and ranges, and comparisons, using Mann-Whitney U test.

## Results

Thirty-eight adults with 22q11DS and thirteen age-matched controls participated in the study. Demographic characteristics are summarized in Table 1. The majority of patients (71%) had the large 3Mb 22q11.2 deletion, and 21% had a history of psychosis at the time of evaluation, similar to what has been described in other large cohorts (McDonald-McGinn et al., 2015).

**Table 1.** General characteristics of participants

|  | Cases | Controls | p value |
| --- | --- | --- | --- |
| n | 38 | 13 |  |
| Male, n (%) | 17 (44.7) | 5 (38.5) | n.s. |
| Female, n (%) | 21 (55.3) | 8 (61.5) |  |
| Median age (years) (range) | 22 (18-50) | 24 (22-33) | n.s. |
| Deletion size 3Mb (32 with known size), n (%) | 27 (71.1) |  |  |
| Psychosis n (%) | 8 (21) | 0 |  |
| Full Scale IQ, median (range) | 71.5(47-93) |  |  |

n.s. not significant

The patients scored significantly lower in olfactory tests, and higher in autonomic dysfunction compared to age-matched controls. In particular, gastrointestinal and pupillomotor concerns in the COMPASS-31 showed the largest difference between cases and controls (Table 2).

**Table 2.**
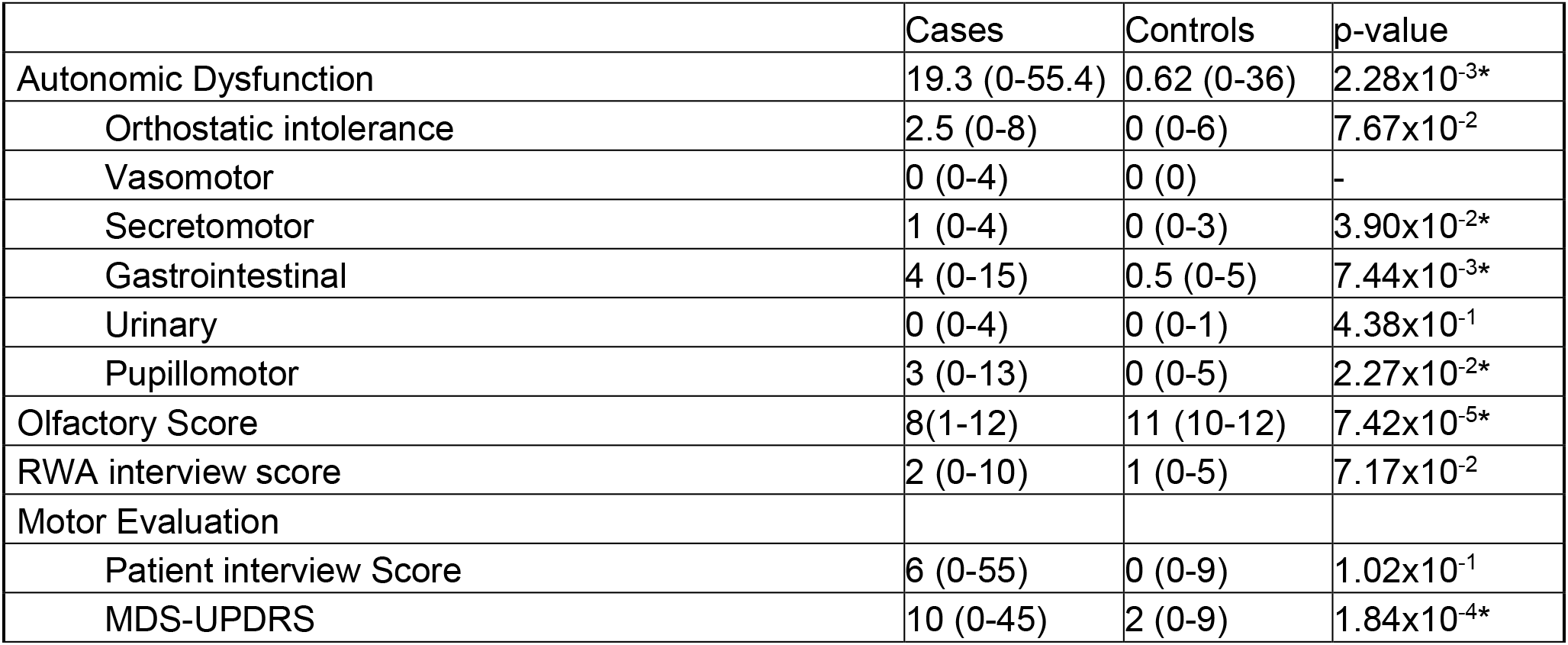
Prodromal PD scales results (* p<0.05). Values correspond to median and range

The motor evaluation showed no statistically significant difference in self-reported motor symptoms of concern between cases and controls, but the cases scored significantly higher in the MDS-UPDRS evaluation, reflecting the presence of mild motor signs (Table 2).

Regarding the sleep evaluation, patients scored higher in RBD questionnaires compared to controls. Home PSG were performed only in cases, and none had findings of REM sleep without atonia, a hallmark of RBD. Detailed findings on these assessments were reported by Mauro et al (Mauro et al., 2022).

To evaluate the integrity of dopaminergic nigro-striatal neurons in the participants, Positron PET/CT imaging was performed using ^18^F-PR04.MZ, a radio ligand with high affinity and selectivity for presynaptic DAT. On average, cases had higher SBR DAT signaling in the striatum than the age-matched controls. Women with 22qDS had higher SBR than men with 22qDS; this gender difference was not seen in controls (data not shown). Of note, two patients with 22qDS, a 23-year-old woman and a 21-year-old man had decreased striatal SBR (60% of control). The results are summarized in table 3.

**Table 3.** DAT signaling by PET/CT with ^18^F-PR04.MZ (* p<0.05)

| SBR ratio in striatum<br>(average left/right) | 22q11.2<br>Mean (SD) | Control<br>Mean<br>(SD) | p-value |
| --- | --- | --- | --- |
| Substantia Nigra | 5.8 (0.9) | 5 (0.6) | $1.23 \times 10^{-2*}$ |
| Caudate Nucleus | 21.8 (4.2) | 18.2 (2.4) | $9.57 \times 10^{-3*}$ |
| Putamen | 23.2 (4.2) | 20.9 (2.9) | $1.96 \times 10^{-2*}$ |

To assess whether the presence of psychosis influenced the prodromal manifestations, we compared these results in persons with the deletion, with (n=8) or without psychosis (N=30). No significant differences were found among the 2 groups (Table 4).

**Table 4.** Subgroup analysis of 22q11.2 patients with or without psychosis (* p<0.05) Values correspond to median and range

|  | With psychosis<br>(n=8)<br>mean (range) | Without psychosis<br>(n=30)<br>mean (range) | p Value |
| --- | --- | --- | --- |
| Autonomic Dysfunction | 21.5 (2.9-30.6) | 18 (0-55.4) | 1.00 |
| Orthostatic intolerance | 2.5 (0-5) | 2.5 (0-7) | 0.95 |
| Vasomotor | 0 (0-4) | 0 (0-4) | 0.38 |
| Secretomotor | 1 (0-3) | 1 (0-4) | 0.58 |
| Gastrointestinal | 4 (0-15) | 3.5 (0-13) | 0.89 |
| Urinary | 0 (0-2) | 0 (0-4) | 0.56 |
| Pupillomotor | 3.5 (0-8) | 2.5 (0-13) | 0.54 |
| Olfactory Score | 8(5-10) | 8 (1-12) | 0.43 |
| RWA interview score | 3 (0-5) | 2 (0-10) | 0.76 |
| Motor Evaluation |  |  |  |
| MDS-UPDRS | 12 (6-45) | 7 (8-18) | 0.28 |
| Patient interview Score | 10.5 (0-39) | 0 (0-41) | 0.19 |

The findings from all these prodromal markers were integrated in the calculation of a likelihood ratio (LR) for prodromal PD, according to Berg et al(Berg et al., 2015). For young adults, such as the participants in this study, a LR of 1000 or above is considered highly suggestive of prodromal PD. One case of the individuals with low SBR fulfilled this criterion, also having a high MDS-UPDRS score and hyposmia.

Finally, we performed exome sequencing of 33 patients with 22qDS, and searched for additional risk factors or modifiers variants in 24 known genes associated with monogenic PD or Parkinsonism. No predicted pathogenic or likely pathogenic variants were found.

## Discussion

To our knowledge, this is the largest study of prodromal PD in adults with 22qDS, including the largest dataset on PET/CT for dopaminergic integrity. Persons with this condition have been recently shown to have a high risk of developing PD ((Butcher et al., 2013), and we sought to identify features that can illustrate the trajectory towards this neurodegenerative disorder and identify individuals in early stages of this progression. In this cross-sectional design, we observed the presence of hyposmia, dysautonomia and mild movement alterations, suggestive of prodromal PD findings in young adults with 22qDS. These findings provide additional evidence, supporting the results reported by Buckley et al (Buckley et al., 2017) and Butcher et al (Butcher et al., 2017b). We found one young person who fulfilled prodromal PD features, and one other participant with reduced DAT signaling on PET-CT.

Except for these two patients, we did not find the more specific markers of higher risk of prodromal PD, namely a decrease in dopaminergic signaling, and RBD/REM sleep without atonia. There were motor alterations, but we found a paradoxical increase in striatal DAT signaling in cases compared to controls. We hypothesize that this increase can be due to decreased dopamine metabolism given haploinsufficiency of the COMT gene, located in the 22qDS region, that encodes for the catechol-O-methyltransferase, as has been shown measuring urine catecholamine metabolites and also by neuroimaging (Boot, Booij, Abeling, et al., 2011; Boot, Booij, Zinkstok, et al., 2011; Boot et al., 2008; Butcher et al., 2017a). This elevated dopaminergic tone has been implicated in the pathogenesis of psychosis, a common manifestation in 22qDS, but it is unknown if it causes a subsequent risk of dopaminergic neuron loss. A role for dopamine autotoxicity has been proposed (Goldstein et al., 2014), but PD risk is not known to be elevated in persons with other hyperdopaminergic states, such as multifactorial psychosis. Alternatively, these 2 phenomena may be unrelated, and other mechanisms altered by the deletion may be the cause of the elevated PD risk.

This study has several limitations. On one hand, the cross-sectional design does not allow to understand the predictive value of these markers in the future progression to PD. In addition, these prodromal markers have not been validated in this 22qDS population, and may be confounded by other factors, such as IQ and the presence of several comorbidities. The small number of participants, a reality in studies of rare disorders, may be underpowered to detect low frequency genetic variants in PD or other genes that may impact risk.

In summary, patients with 22qDS at high risk for PD show findings suggestive of prodromal PD, but follow-up of these patients and further longitudinal studies are relevant to understand the development of and progression to PD in this high-risk population.

## Data Availability

All data produced in the present study are available upon reasonable request to the authors

## References

Berg, D., Postuma, R. B., Adler, C. H., Bloem, B. R., Chan, P., Dubois, B., Gasser, T., Goetz, C. G., Halliday, G., Joseph, L., Lang, A. E., Liepelt-Scarfone, I., Litvan, I., Marek, K., Obeso, J., Oertel, W., Olanow, C. W., Poewe, W., Stern, M., & Deuschl, G. (2015). MDS research criteria for prodromal Parkinson’s disease. Movement Disorders : Official Journal of the Movement Disorder Society, 30(12), 1600–1611. https://doi.org/10.1002/mds.26431

Blagojevic, C., Heung, T., Theriault, M., Tomita-Mitchell, A., Chakraborty, P., Kernohan, K., Bulman, D. E., & Bassett, A. S. (2021). Estimate of the contemporary live-birth prevalence of recurrent 22q11.2 deletions: a cross-sectional analysis from population-based newborn screening. CMAJ Open, 9(3), E802–E809. https://doi.org/10.9778/CMAJO.20200294

Booij, J., Van Amelsvoort, T., & Boot, E. (2010). Co-occurrence of early-onset Parkinson disease and 22q11.2 deletion syndrome: Potential role for dopamine transporter imaging. In American Journal of Medical Genetics, Part A (Vol. 152, Issue 11, pp.2937–2938). Am J Med Genet A. https://doi.org/10.1002/ajmg.a.33665

Boot, E., Booij, J., Abeling, N., Meijer, J., da Silva Alves, F., Zinkstok, J., Baas, F., Linszen, D., & van Amelsvoort, T. (2011). Dopamine metabolism in adults with 22q11 deletion syndrome, with and without schizophrenia--relationship with COMT Val^108^/^158^Met polymorphism, gender and symptomatology. Journal of Psychopharmacology (Oxford, England), 25(7), 888–895. https://doi.org/10.1177/0269881111400644

Boot, E., Booij, J., Zinkstok, J., Abeling, N., de Haan, L., Baas, F., Linszen, D., & van Amelsvoort, T. (2008). Disrupted dopaminergic neurotransmission in 22q11 deletion syndrome. Neuropsychopharmacology : Official Publication of the American College of Neuropsychopharmacology, 33(6), 1252–1258. https://doi.org/10.1038/sj.npp.1301508

Boot, E., Booij, J., Zinkstok, J. R., Baas, F., Swillen, A., Owen, M. J., Murphy, D. G., Murphy, K. C., Linszen, D. H., & van Amelsvoort, T. A. (2011). COMT Val(158) met genotype and striatal D(2/3) receptor binding in adults with 22q11 deletion syndrome. Synapse (New York, N.Y.), 65(9), 967–970. https://doi.org/10.1002/syn.20932

Braak, H., Bohl, J. R., Müller, C. M., Rüb, U., de Vos, R. A. I., & Del Tredici, K. (2006). Stanley Fahn Lecture 2005: The staging procedure for the inclusion body pathology associated with sporadic Parkinson’s disease reconsidered. Movement Disorders : Official Journal of the Movement Disorder Society, 21(12), 2042–2051. https://doi.org/10.1002/mds.21065

Braak, H., Del Tredici, K., Rüb, U., de Vos, R. A. I., Jansen Steur, E. N. H., & Braak, E. (2003). Staging of brain pathology related to sporadic Parkinson’s disease. Neurobiology of Aging, 24(2), 197–211.

Buckley, E., Siddique, A., & McNeill, A. (2017). Hyposmia, symptoms of rapid eye movement sleep behavior disorder, and parkinsonian motor signs suggest prodromal neurodegeneration in 22q11 deletion syndrome. NeuroReport, 28(11), 677–681. https://doi.org/10.1097/WNR.0000000000000815

Butcher, N. J., Kiehl, T.-R., Hazrati, L.-N., Chow, E. W. C., Rogaeva, E., Lang, A. E., & Bassett, A. S. (2013). Association between early-onset Parkinson disease and 22q11.2 deletion syndrome: identification of a novel genetic form of Parkinson disease and its clinical implications. JAMA Neurology, 70(11), 1359–1366. https://doi.org/10.1001/jamaneurol.2013.3646

Butcher, N. J., Marras, C., Pondal, M., Rusjan, P., Boot, E., Christopher, L., Repetto, G. M., Fritsch, R., Chow, E. W. C., Masellis, M., Strafella, A. P., Lang, A. E., & Bassett, A. S. (2017a). Neuroimaging and clinical features in adults with a 22q11.2 deletion at risk of Parkinson’s disease. Brain, 140(5), 1371–1383. https://doi.org/10.1093/brain/awx053

Butcher, N. J., Marras, C., Pondal, M., Rusjan, P., Boot, E., Christopher, L., Repetto, G. M., Fritsch, R., Chow, E. W. C., Masellis, M., Strafella, A. P., Lang, A. E., & Bassett, A. S. (2017b). Neuroimaging and clinical features in adults with a 22q11.2 deletion at risk of Parkinson’s disease. Brain, 140(5), 1371–1383. https://doi.org/10.1093/brain/awx053

Ferrando, L., Soto, M., Bobes, J., Soto, O., Franco, L., & Gubert, J. (1998). M.I.N.I. Mini International Neuropsychiatric Interview. Versión en Español 5.0.0 DSM-IV. Instituto IAP.

Frauscher, B., & Högl, B. (2015). Quality Control for Diagnosis of REM sleep behavior disorder: Criteria, Questionnaires, Video and Polysomnography. In A. Videnovic and B Högl (Ed.), Disorders of Sleep and Circadian Rythm in Parksinson’s Disease (pp. 145–157). Springer-Verlag. https://doi.org/10.1007/978-3-7091-1631-9_11

Genome Aggregation Database (gnomAD). (n.d.). gnomAD. Retrieved June 10, 2020, from https://gnomad.broadinstitute.org/

Goetz, C. G., Tilley, B. C., Shaftman, S. R., Stebbins, G. T., Fahn, S., Martinez-Martin, P., Poewe, W., Sampaio, C., Stern, M. B., Dodel, R., Dubois, B., Holloway, R., Jankovic, J., Kulisevsky, J., Lang, A. E., Lees, A., Leurgans, S., LeWitt, P. A., Nyenhuis, D., … Zweig, R. M. (2008). Movement Disorder Society-Sponsored Revision of the Unified Parkinson’s Disease Rating Scale (MDS-UPDRS): Scale presentation and clinimetric testing results. Movement Disorders, 23(15), 2129–2170. https://doi.org/10.1002/mds.22340

Goldstein, D. S., Kopin, I. J., & Sharabi, Y. (2014). Catecholamine autotoxicity. Implications for pharmacology and therapeutics of Parkinson disease and related disorders. Pharmacology & Therapeutics, 144(3), 268–282. https://doi.org/10.1016/j.pharmthera.2014.06.006

Harris, P. A., Taylor, R., Thielke, R., Payne, J., Gonzalez, N., & Conde, J. G. (2009). Research electronic data capture (REDCap)--a metadata-driven methodology and workflow process for providing translational research informatics support. Journal of Biomedical Informatics, 42(2), 377–381. https://doi.org/10.1016/J.JBI.2008.08.010

Iber, C., Ancoli-Israel, S., Chesson, A. J., Quan, S., & Medicine, for the A. A. of S. (2007). The AASM manual for the scoring of sleep and associated events: rules, terminology and technical specifications.The AASM manual for the scoring of sleep and associated events: rules, terminology and technical specifications. American Academy of Sleep Medicine.

Juri, C., Kramer, V., Riss, P. J., Soza-Ried, C., Haeger, A., Pruzzo, R., Rösch, F., Amaral, H., & Chana-Cuevas, P. (2021). PR04.MZ PET/CT Imaging for Evaluation of Nigrostriatal Neuron Integrity in Patients with Parkinson Disease. Clinical Nuclear Medicine, 46(2), 119–124. https://doi.org/10.1097/RLU.0000000000003430

Kircher, M., Witten, D. M., Jain, P., O’roak, B. J., Cooper, G. M., & Shendure, J. (2014). A general framework for estimating the relative pathogenicity of human genetic variants. Nature Genetics 2014 46:3, 46(3), 310–315. https://doi.org/10.1038/ng.2892

Kramer, V., Juri, C., Riss, P. J., Pruzzo, R., Soza-Ried, C., Flores, J., Hurtado, A., Rösch, F., Chana-Cuevas, P., & Amaral, H. (2020). Pharmacokinetic evaluation of [18F]PR04.MZ for PET/CT imaging and quantification of dopamine transporters in the human brain. European Journal of Nuclear Medicine and Molecular Imaging, 47(8), 1927–1937. https://doi.org/10.1007/s00259-019-04594-z

Lehnert, W., Riss, P. J., Hurtado de Mendoza, A., Lopez, S., Fernandez, G., Ilheu, M., Amaral, H., & Kramer, V. (2022). Whole-body biodistribution and radiation dosimetry of [18F]PR04.MZ: a new PET radiotracer for clinical management of patients with movement disorders. EJNMMI Research, 12(1), 1. https://doi.org/10.1186/s13550-021-00873-9

Mauro, J., DIaz, M., Córdova, T., Villanueva, K., Cáceres, T., Bassi, A., Fritsch, R., Repetto, G. M., & Ocampo-Garcés, A. (2022). Analysis of REM sleep without atonia in 22q11.2 deletion syndrome determined by domiciliary polysomnography: A cross sectional study. Sleep, 45(2). https://doi.org/10.1093/sleep/zsab300

McDonald-McGinn, D. M., Sullivan, K. E., Marino, B., Philip, N., Swillen, A., Vorstman, J. A. S., Zackai, E. H., Emanuel, B. S., Vermeesch, J. R., Morrow, B. E., Scambler, P. J., & Bassett, A. S. (2015). 22Q11.2 Deletion Syndrome. Nature Reviews Disease Primers, November, 15071. https://doi.org/10.1038/nrdp.2015.71

Mok, K. Y., Sheerin, U., Simón-Sánchez, J., Salaka, A., Chester, L., Escott-Price, V., Mantripragada, K., Doherty, K. M., Noyce, A. J., Mencacci, N. E., Lubbe, S. J., Williams-Gray, C. H., Barker, R. A., van Dijk, K. D., Berendse, H. W., Heutink, P., Corvol, J.-C., Cormier, F., Lesage, S., … Wood, N. W. (2016). Deletions at 22q11.2 in idiopathic Parkinson’s disease: a combined analysis of genome-wide association data. The Lancet. Neurology. https://doi.org/10.1016/S1474-4422(16)00071-5

Oskarsdóttir, S., Vujic, M., & Fasth, A. (2004). Incidence and prevalence of the 22q11 deletion syndrome: a population-based study in Western Sweden. Archives of Disease in Childhood, 89(2), 148–151.

Panamonta, V., Wichajarn, K., Chaikitpinyo, A., Panamonta, M., Pradubwong, S., & Chowchuen, B. (2016). Birth Prevalence of Chromosome 22q11.2 Deletion Syndrome: A Systematic Review of Population-Based Studies. Journal of the Medical Association of Thailand = Chotmaihet Thangphaet, 99, S187–S193.

Pollard, R., Hannan, M., Tanabe, J., & Berman, B. D. (2016). Early-onset Parkinson disease leading to diagnosis of 22q11.2 deletion syndrome. Parkinsonism & Related Disorders, 25, 110–111. https://doi.org/10.1016/j.parkreldis.2016.01.027

Postuma, R. B., Arnulf, I., Hogl, B., Iranzo, A., Miyamoto, T., Dauvilliers, Y., Oertel, W. Ju,, Y. el, Puligheddu, M., Jennum, P., Pelletier, A., Wolfson, C., Leu-Semenescu, S., Frauscher, B., Miyamoto, M., Cochen De Cock, V., Unger, M. M., Stiasny-Kolster, K., Livia Fantini, M., & Montplaisir, J. Y. (2012). A single-question screen for rapid eye movement sleep behavior disorder: A multicenter validation study. Movement Disorders, 27(7), 913–916. https://doi.org/10.1002/mds.25037

Postuma, R. B., Berg, D., Stern, M., Poewe, W., Olanow, C. W., Oertel, W., Obeso, J., Marek, K., Litvan, I., Lang, A. E., Halliday, G., Goetz, C. G., Gasser, T., Dubois, B., Chan, P., Bloem, B. R., Adler, C. H., & Deuschl, G. (2015). MDS clinical diagnostic criteria for Parkinson’s disease. Movement Disorders : Official Journal of the Movement Disorder Society, 30(12), 1591–1601. https://doi.org/10.1002/mds.26424

Postuma, R. B., Lang, A. E., Gagnon, J. F., Pelletier, A., & Montplaisir, J. Y. (2012). How does parkinsonism start? Prodromal parkinsonism motor changes in idiopathic REM sleep behaviour disorder. Brain : A Journal of Neurology, 135(Pt 6), 1860–1870. https://doi.org/10.1093/brain/aws093

Rehman, A. F., Dhamija, R., Williams, E. S., & Barrett, M. J. (2015). 22q11.2 deletion syndrome presenting with early-onset Parkinson’s disease. Movement Disorders : Official Journal of the Movement Disorder Society, 30(9), 1289–1290. https://doi.org/10.1002/mds.26305

Romanos, M., Schecklmann, M., Kraus, K., Fallgatter, A. J., Warnke, A., Lesch, K.-P., & Gerlach, M. (2011). Olfactory deficits in deletion syndrome 22q11.2. Schizophrenia Research, 129(2–3), 220–221. https://doi.org/10.1016/j.schres.2010.12.015

Rosas, R., Tenorio, M., Pizarro, M., Cumsille, P., Bosch, A., Arancibia, S., Carmona-Halty, M., Pérez-Salas, C., Pino, E., Vizcarra, B., & Zapata-Sepúlveda, P. (2014). Estandarización de la Escala Wechsler de Inteligencia Para Adultos-Cuarta Edición en Chile. Psykhe (Santiago), 23(1), 1–18. https://doi.org/10.7764/psykhe.23.1.529

Sletten, D. M., Suarez, G. A., Low, P. A., Mandrekar, J., & Singer, W. (2012). COMPASS 31: a refined and abbreviated Composite Autonomic Symptom Score. Mayo Clinic Proceedings, 87(12), 1196–1201. https://doi.org/10.1016/j.mayocp.2012.10.013

Sobin, C., Kiley-Brabeck, K., Dale, K., Monk, S. H., Khuri, J., & Karayiorgou, M. (2006). Olfactory disorder in children with 22q11 deletion syndrome. Pediatrics, 118(3), e697–703. https://doi.org/10.1542/peds.2005-3114

Stiasny-Kolster, K., Mayer, G., Schäfer, S., Möller, J. C., Heinzel-Gutenbrunner, M., & Oertel, W. H. (2007). The REM sleep behavior disorder screening questionnaire -A new diagnostic instrument. Movement Disorders, 22(16), 2386–2393. https://doi.org/10.1002/mds.21740

Zaleski, C., Bassett, A. S., Tam, K., Shugar, A. L., Chow, E. W. C., & McPherson, E. (2009). The co-occurrence of early onset Parkinson disease and 22q11.2 deletion syndrome. American Journal of Medical Genetics. Part A, 149A(3), 525–528. https://doi.org/10.1002/ajmg.a.32650

